# High throughput screening of nuclear receptors identifies NR4A1, a novel tumor suppressor with potential as a therapeutic target in gallbladder cancer

**DOI:** 10.1101/2024.03.13.24304218

**Authors:** Sajib Kumar Sarkar, Rashmi Minocha, Prasenjit Das, Nihar Ranjan Dash, Ruby Dhar, Deepak Kumar, Vinay Kumar Kapoor, Ratnakar Shukla, Subhradip Karmakar

## Abstract

**Introduction:** Gallbladder cancer (GBC) is one of the most common cancer of the hepato-biliary tract, with a strikingly variable incidence and prevalence across different regions of the world. The Indo-Gangetic belt in Northern India is reported to have one of the highest incidences of about 21/100,000. GBC usually goes unnoticed due to the lack of any early symptoms with two third of GBC cases present late at inoperable stages and have very limited treatment options. Nuclear receptors, a family of 48 members are ligand dependent transcription factors. They are of particular interest in cancer research because of their established role in cancer pathogenesis and their excellent druggability that makes them a suitable therapeutic target.

**Methodology:** mRNA expression 48 nuclear receptors were assessed in GBC tissue samples (n=13) and chronic cholecystitis tissue samples by Nanostring nCounter. The screening identified orphan receptor NR4A1 to be significantly downregulated in GBC. Western blot were performed to further validate the same. We next interrogated the above findings in 2 different gallbladder cancer cell lines, the highly invasive NOZ and the non-invasive TGBC24TKB. In order to investigate the role of NR4A1 in GBC pathogenesis, NOZ cells were treated with cytosporone B (10µM for 24hours) an agonist of NR4A1. On the other hand NR4A1 was knocked down in TGBC24TKB by siRNA. Expression of different markers of proliferation, invasion and epithelial mesenchymal transition was assessed by qPCR. Cell cycle analysis was done using flow cytometry.

**Results:** NR4A1 was one of the top differentially expressed (down regulated) nuclear receptors in GBC both in RNA and protein level. Similar finding was observed in highly invasive cell line NOZ in comparison to TGBC24TKB. Cytosporone B treatment led to upregulation of NR4A1, which resulted in reduction of cell migration as evident by delayed wound healing, reduction in invasion with an increase in G0/G1 populations implying a growth arrest. NR4A1 knockdown in TGBC24TKB lead to reduction in G0/G1 fraction and also increase in proliferation markers like mki67.

**Conclusion:** NR4A1 in our study acts as a tumor suppressor, loss of which seems to provide a growth and survival advantage to GBC cells. NR4A1 activation by agonist reduced cell proliferation and invasion. We therefore propose NR4A1 as a novel biomarker in GBC with its loss associated with overall poor outcome. Hence its agonists may emerge as a potential candidate for neo-adjuvant therapy for advanced gallbladder cancer.

## Introduction

Gallbladder cancer (GBC) is the most common malignancy of the biliary tract worldwide(1). Incidence of GBC exhibits significant geographical and ethnic variation. Bolivia is the country with highest incidence of GBC (∼14/lakh). In India the incidence ranges from scarce in the southern part to very high (upto 22/lakh) in few districts of North-eastern India(2).

Global cancer statistics 2020: GLOBOCAN projections estimate that by 2025, the number of patients with GBC in India will reach approximately 9.8 and 11.2 per cent among men and women, respectively, across all patients worldwide. Also if no specific intervention is implemented to ameliorate GBC risk factors in India, it will replace Chile as the country with the highest GBC incidence and mortality(3). Therefore, all possible measures should be urgently undertaken to reduce the incidence of GBC in India, especially in the northern region.

The Indian Council of Medical Research (ICMR) identified several risk factors for GBC including ethnicity, gender, age, gallstones, chronic inflammation, genetic factors, gallbladder polyps and lifestyle(4). However, specific risk factors for GBC in the northern region of India have not yet been identified. The ICMR has provided information on obesity control and diabetes, adopting a healthy diet rich in fruits and vegetables and exercising regularly as preventive measures for the development of GBC. The presence of gallstones is considered to be the most important risk factor. Only 1-3 per cent of cholelithiasis patients develop GBC(5), and the frequency of Indian GBC patients with gallstones is reportedly 70-90 per cent(6). This suggests that factors in addition to gallstones and molecular players involved in GBC development are yet to be identified.

The NR superfamily consists of 48 members having similar modular structure. These ligand dependent transcription factors are of particular interest in cancer research because of their regulatory role in cellular metabolism, proliferation, differentiation etc as well as their excellent druggablity(7). Estrogen and progesterone receptors in breast cancer, androgen receptors in prostate cancer, retinoic acid receptor in acute myelogenous leukemia are well studied(8–10). Also, bile contains high concentration of cholesterol and numerous lipophilic cholesterol derivatives which gets concentrated further in gallbladder. These molecules are ligands/ potential ligands of intracellular nuclear receptors of GB epithelium(11). Hence, we hypothesize that dysregulation of nuclear receptor signaling may contribute to the pathogenesis of GBC.

Our approach was to identify differentially expressed nuclear receptors in GBC followed by exploring the effect of manipulation of these receptors in gallbladder cancer cell lines.

## Results

### 1. NR4A1 is downregulated in GBC (Tissue)

To identify differentially expressed nuclear receptors, Nanostring nCounter technique was used with a customized panel for nuclear receptors (48 targets and 6 housekeeping, Supplementary Table1). Nanostring is a high throughput probe-based transcript counting technique (molecular barcoding) which employs no intermediate amplification step unlike RNA sequencing. Thus, it provides the actual quantity of the transcripts under investigation(12).

Analysis of nuclear receptors superfamily (48 members) revealed that members of nuclear receptor family 4 NR4A1 and NR4A3 were amongst the highest expressed nuclear receptors in control gallbladder (GB) tissue (based on transcript count). Both were significantly downregulated in GBC (n=13) (Fig. 1a and S1a). NR4A2 was downregulated too but was not statistically significant (Fig. S1b).

**Figure 1:**
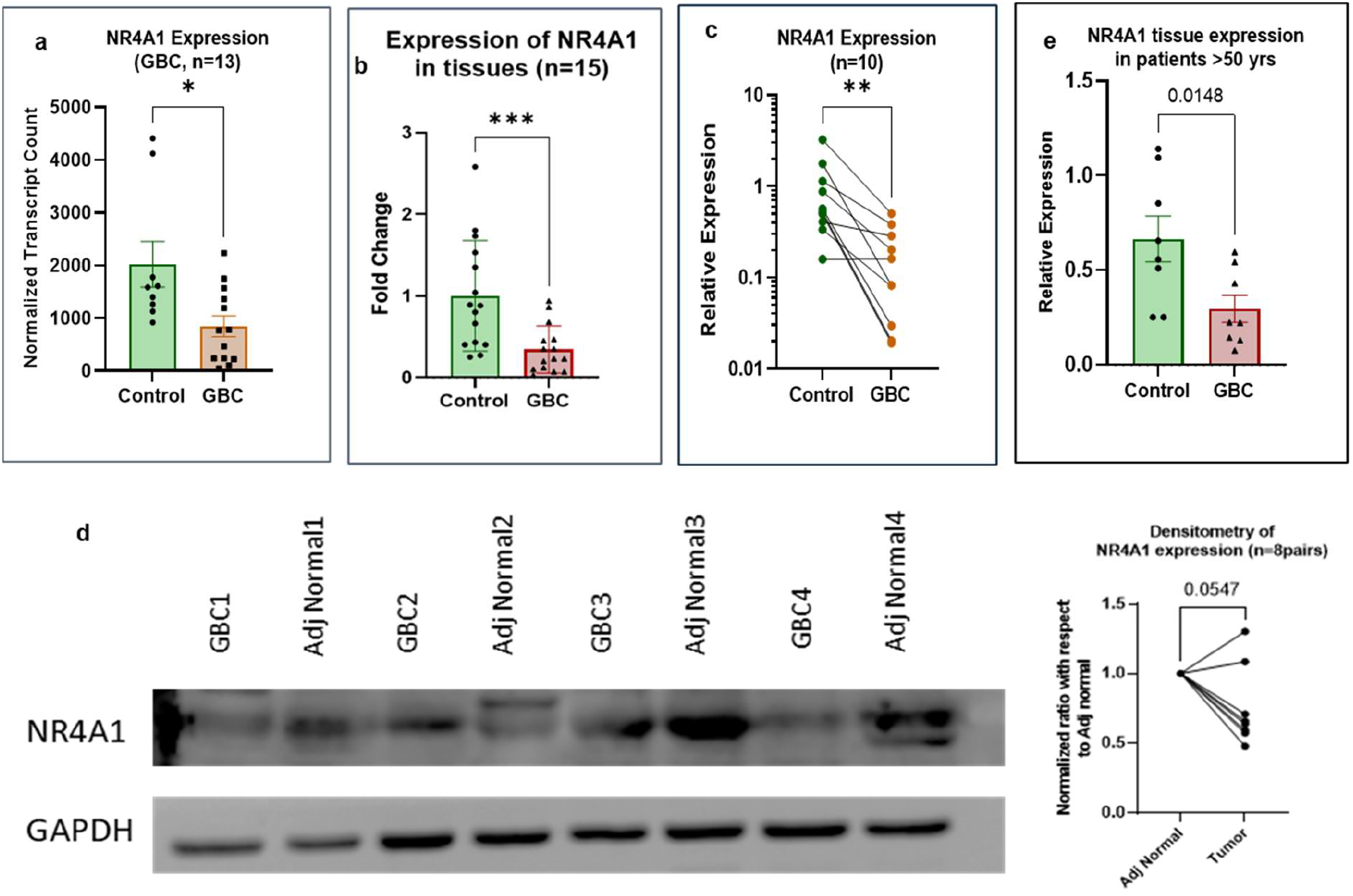
Expression of NR4A1 in Gallbladder cancer. Tissues were lysed by trizol and RNA isolation was done by DNase digestion and column purification. For protein isolation RIPA lysis buffer was used. a,b) Expression of NR4A1 in GBC compared with unmatched controls by Nanostring nCounter and qPCR respectively. c) NR4A1 expression in GB tumor compared to adjacent normal GB tissue (matched). d) NR4A1 protein level expression in GB tumor and matched Adjacent normal GB tissue (Adj Normal) along with densitometric analysis. e) Expression of NR4A1 in patients with age >50 years. [Mann-Whitney U test was done to analyze statistical significance]

To further verify, expression of NR4A1 in more GBC tissues (n=15) was analyzed by qPCR and was compared to unrelated non-malignant GB tissue (chronic cholecystitis) (Fig.1b). The mean expression in GBC was found to be one third of that of the non-malignant GB tissue and the difference was statistically significant (Mann-Whitney *p* value= 0.0009). Additionally, we also found significant downregulation of NR4A1 in GBC tumor tissue compared to paired adjacent normal GB tissue of the same patient (n=10, Wilcoxon test *p* value= 0.0039, Fig. 1c). Protein level of the receptor too reflected the same when NR4A1 in tumor tissue was compared with that of adjacent normal GB tissue (Fig, 1d). Downregulation of NR4A1 was seen both in male and female GBC. We found no significant sex bias in the expression pattern of NR4A1. Although the NR4A1 downregulation significantly more evident in patients age more than 50 years (Fig. 1e). TCGA database mining reveals significant downregulation of NR4A1 in most of the cancers while overexpression was observed in some eg. Pancreatic adenocarcinoma, Lymphoid malignancies etc (Fig. S1d).

### 2. Expression of NR4A1 is downregulated in invasive GBC cell lines

Of 3 different GBC cell lines NOZ and G415 were found to be highly invasive (by trans-well Matrigel invasion assay) (Fig.2a), having higher proportion of cells in S and G2M phase of cell cycle (by PI staining) (Fig.2b), faster wound closure when compared to TGBC24TKB which was non-invasive. Also, NOZ and G415 expressed more mesenchymal markers eg N-Cadherin, vimentin while TGBC24TKB highly expressed both E-Cadherin and N-Cadherin. Thus, TGBC24TKB is non-invasive and with more epithelial markers represent a well differentiated form of GBC. NOZ and G415 represents a form of invasive GBC. NR4A1 was found to be downregulated in aggressive cell lines G415 and NOZ in comparison to non-invasive cell line TGBC24TKB (Fig. 2c).

**Figure 2:**
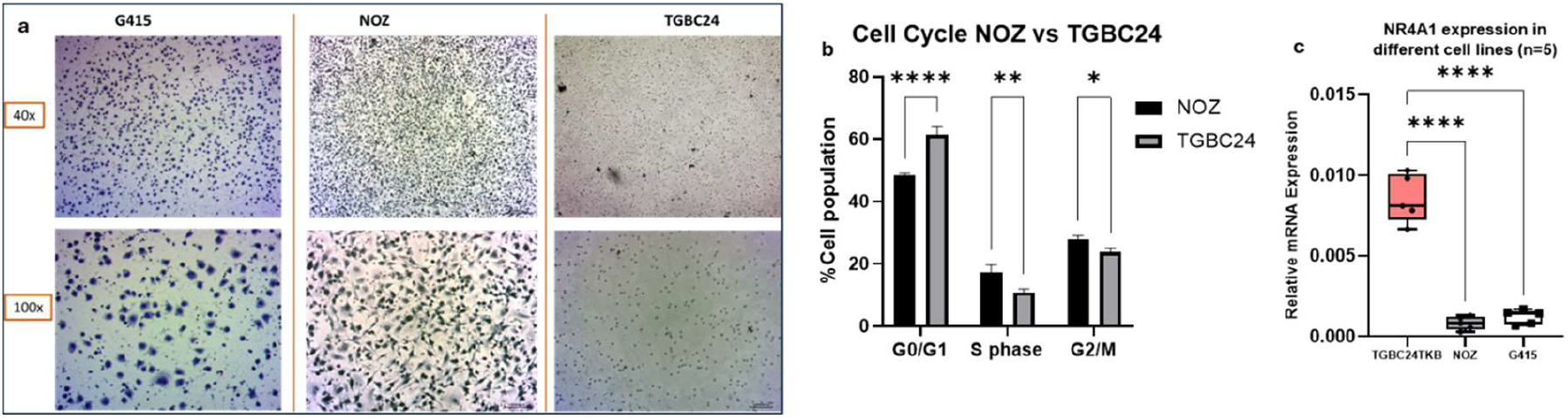
Characterization of GBC cell lines. a) Transwell migration assay with matrigel coated insert shows the migrated cells on the undersurface of the membrane (stained with crystal violet). b) Propidium iodide staining was used to determine the DNA content of cells analyzed by flowcytometry. c) mRNA expression of NR4A1 in different cell lines by qPCR with GAPDH as a housekeeping gene. [Mann-Whitney U test or Kruskal Wallis test was done to analyze statistical significance]

### 3. Cytosporone B stimulation reduced proliferative potential and invasion of invasive GBC cell line

Cytosporone B (CSNB) is a natural ligand of NR4A1. Treatment with CSNB leads to activation of NR4A1 leading to induction of NR4A1 itself (NR4A1 is induced by NR4A1)(13). We were able to induce NR4A1 in low expressing cell line NOZ as evident by qPCR (Fig.3a). To start with, MTT assay was done to identify cytotoxic/inhibitory effects of CSNB. A dose of 10uM was selected based on the findings i.e. much less than the IC_50_ calculated (60uM) (Fig.S2a). GBC cell lines were treated with 10uM of CSNB and expression of NR4A1 was evaluated. It was observed that NR4A1 transcript level started increasing 4 hours following treatment, peaked at 12 hours and maintained an elevated level till 48 hours (Fig.S2b). Analyzing the outcome, a duration of 24 hours was decided for most of the CSNB treatment.

Immunofluorescence showed that NR4A1 when unbound was chiefly localized in cytosol. On treatment with CSNB there was nuclear translocation of NR4A1 evident at 1 hour post treatment. The receptor is shuttled back to cytosol eventually which is evident at 24 hours post treatment (Fig.S2c). Thus, we have validated CSNB as a ligand of NR4A1 in GBC cell line NOZ.

NR4A1 activation with CSNB led downregulation of proliferation markers like Ki67, PCNA (Fig. 3b,c). Also, there was upregulation of E-Cadherin with concomitant downregulation of N-Cadherin suggesting reverse EMT (Fig. 3d,e).

**Figure 3:**
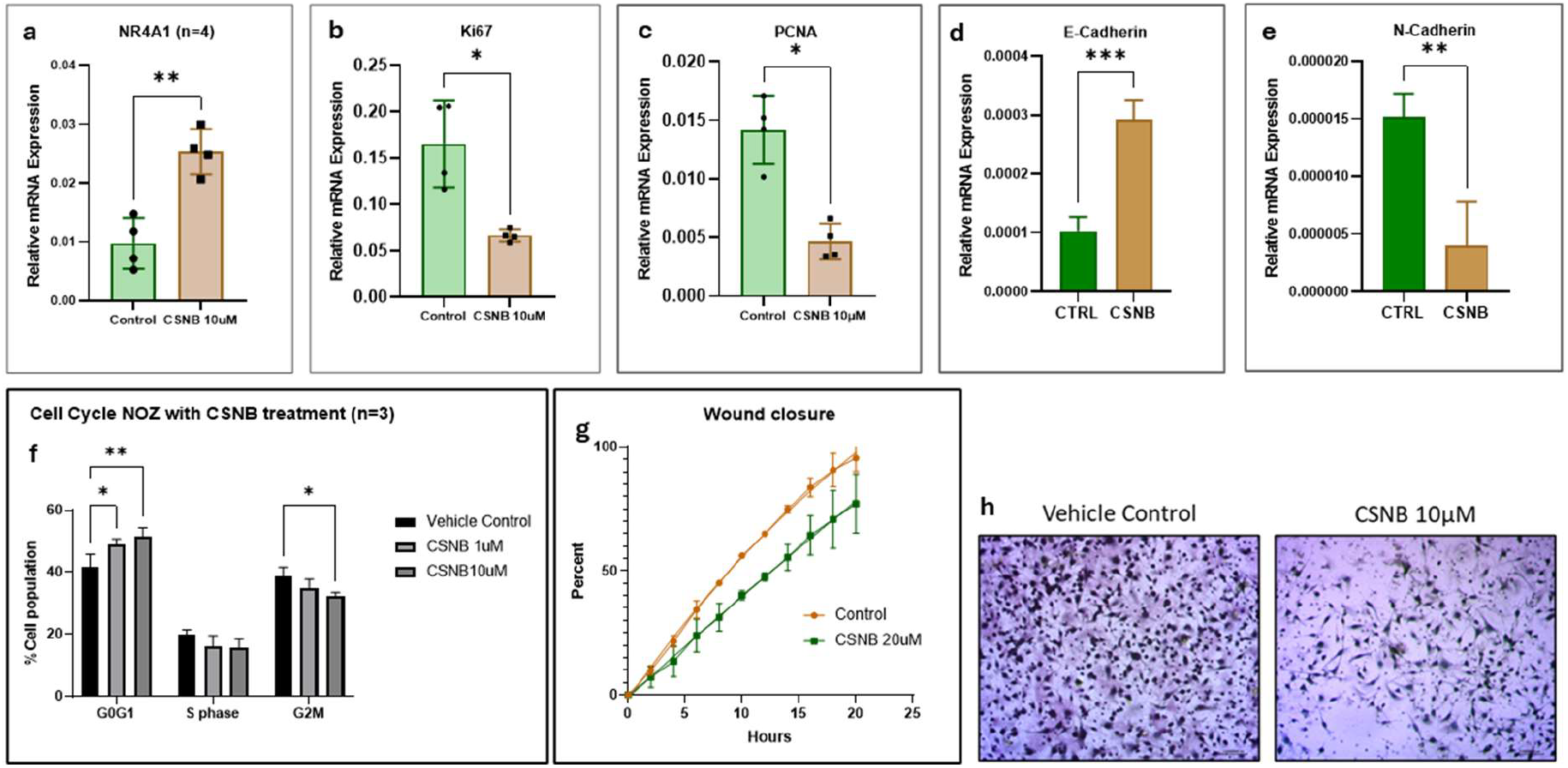
Effect of Cytosporone B treatment on GBC cell line NOZ. a) Upregulation of NR4A1 is indicative of CSNB mediated activation of NR4A1. b-e) expression of different proliferation and EMT markers following CSNB treatment. f)Cell cycle analysis, g)migration analysis by wound healing assay and h) invasion analysis by transwell migration assay following the same treatment. [Mann-Whitney U test or Kruskal Wallis test was done to analyze statistical significance]

Cell cycle analysis suggested significant increase in G_0_/G_1_ fraction while reduction in G_2_/M phase fraction following CSNB treatment (Fig. 3f). There was no significant change in S fraction. We also observed that the effect on cell cycle was dose dependent as there was significant increase in G_0_/G_1_ fraction as dose of CSNB was increased from 1 to 10 µM. This implies a possible G1/S blockade following CSNB treatment.

Wound healing was delayed by CSNB treatment. Vehicle treated wound healed almost completely by 18 hours while CSNB treated wound 1/3^rd^ area was left (Fig. 3g). Transwell Matrigel invasion in NOZ was also reduced by CSNB. Invasion decreased by 30% at 18 hours (Fig. 3h).

### 4. Knockdown of NR4A1 increases proliferation potential and promotes EMT

Knockdown of NR4A1 was achieved by treating cell lines with specific siRNA (20nM) and was verified by qPCR (Fig. 4a). Expression of the other two members of the family eg. NR4A2 and NR4A3 was not altered significantly (Fig. S3a,b).

**Figure 4:**
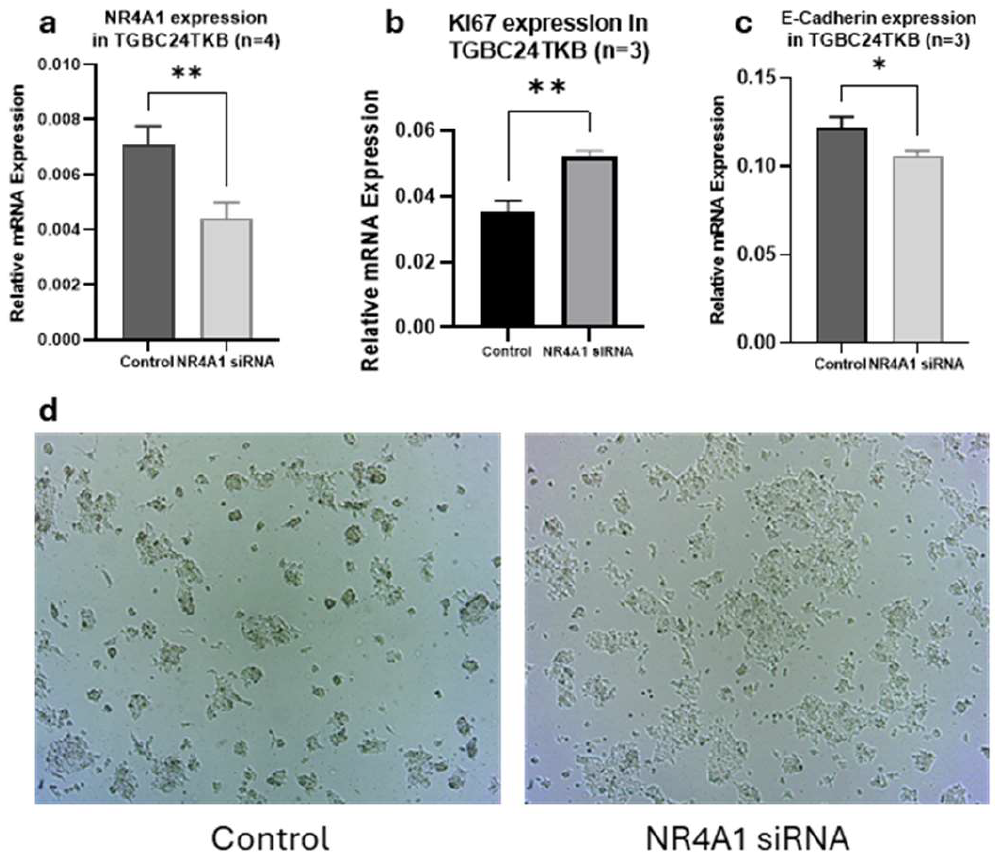
Effect of knocking down NR4A1 in GBC cell line TGBC24TKB by siRNA. a)Knockdown was confirmed by downregulation of NR4A1 mRNA. b,c) expression of proliferation and EMT markers following knockdown. d) Phenotypically proliferation and confluence of TGBC24TKB increased following NR4A1 knockdown.

In TGBC24TKB larger cell colonies and higher confluence was observed microscopically post knockdown of NR4A1 (Fig. 4d). At mRNA level ki67 a proliferation marker expression was increased while expression of E-Cadherin was decreased (Fig. 4b,c). We found no significant difference in expression of PCNA or N-Cadherin or MMP2.

## Discussion

NR4A1 a member of the nuclear receptor superfamily has no known physiological ligand thus known as an orphan receptor. Major regulation of its function is by regulation of its expression. Growth factors, prostaglandins, calcium, cytokines and neurotransmitters are known to induce the expression(14). Additionally, it undergoes different post translational modifications like phosphorylation, acetylation and SUMOylation which further tunes its stability and functionality. NR4A1 exerts its function by binding to response elements as monomer, homo or heterodimer (with RXR) and regulating expression of genes involved in different cellular processes like energy metabolism, steroidogenesis, proliferation, differentiation etc(15). Vast majority of studies on NR4A1 have established its role as a master regulator of cellular metabolism. Physiologically, it upregulates GLUT4 and enhances glucose utilization by skeletal muscle cells by increasing the expression of hexokinase and phosphofructokinase, the regulatory enzymes of glycolysis(16). It is thus considered a promising therapeutic target for metabolic syndromes. Observing its regulatory role in metabolism, NR4A1 has been studied in relation to different cancers too. Interestingly, the overall reported role of NR4A1 in cancer is paradoxical in the literature. It has been reported to have tumor suppressor role in cancers like AML(17), prostate cancer(18), triple negative breast cancer(19). NR4A1 downregulation was found in breast cancer with higher grades. Metastatic disease was mostly NR4A1 negative(20). In hepatocellular carcinoma, NR4A1 promotes gluconeogenesis by stabilizing phosphoenol pyruvate carboxykinase1 (PEPCK1). This leads to inhibition of glycolysis, depletion of ATP and arrest of cell proliferation. Silencing of NR4A1 possibly by promoter hypermethylation leads to severe disease and poor prognosis in HCC(21). On the other hand, several studies have reported NR4A1 as an oncogenic driver. It is upregulated in cancer of lung(22), colon(23) and pancreas(24). It stabilizes HIF1a and promotes its transcriptional activity which supports the tumor in hypoxic environment and set ground for metastasis(25). Thus, it is clear that the impact of NR4A1 is complex and cell dependent.

In current study, we have shown that expression of NR4A1 is downregulated in GBC. Existing expression database supports our finding (TCGA). It also supports tumor suppressor role of NR4A1 as significant downregulation of this receptors occur in most of the cancers except for pancreatic adenocarcinoma, acute myeloid leukemia and thymoma which further supports complex cell specific function of NR4A1.

The two gallbladder cancer cell lines NOZ and TGBC24TKB used for this study were validated by STR profiling (Supplementary data Figure S1). We further characterized the cell lines based on their migration and invasion properties. NOZ is a highly invasive cell line with high migratory property evident by rapid wound healing. On the other hand, TGBC24TKB did not invade the Matrigel matrix and had limited migratory property as evident by non-healing of wound created (Supplementary data Figure S2). We found that NR4A1 was significantly downregulated in highly invasive cell line NOZ in comparison to TGBC24TKB.

In NOZ Cytosporone B activated NR4A1 as evident by nuclear translocation of the receptor following treatment and induction of NR4A1 itself. CSNB has been established as a ligand for NR4A1 previously and is known to cause nuclear translocation and activation of transcriptional activity. NR4A1 itself is induced by its transactivation and has been used as validation of treatment in studies(26). CSNB treatment also resulted in increased G0G1 fraction in NOZ. Also rate of migration was reduced supported by delayed wound closure suggesting anti proliferative tumor suppressor role of NR4A1.

In TGBC24TKB, a non-invasive cell line, NR4A1 knockdown led to increased expression of proliferation marker like Ki67 and reduced expression of epithelial marker E-Cadherin. Increased proliferation rate was microscopically evident by higher confluence following the knockdown.

NR4A1 seems to have tumor suppressor role in GBC. As it is downregulated in GBC, Induction of the receptor with a ligand may serve as a potential adjuvant therapeutic modality.

## Data Availability

All data produced in the present study are available upon reasonable request to the authors

## Acknowledgement

The authors acknowledge Indian Council of Medical Research (ICMR) for funding the study.

## Methodology

### Ethics approval

Appropriate approval and certification by Institute Ethics Committee, All India Institute of Medical Sciences, New Delhi was obtained prior to the start of the study.

#### 1. Tissue collection

GB Tumor tissue was collected from cases of GBC undergoing extended cholecystectomy. In 5 GBC cases adjacent normal GB tissue were also collected and these were used as paired control for those GBC tumor tissue.

Control GB tissue was collected from patients of chronic cholecystitis undergoing simple cholecystectomy. Informed consent was taken for each participant.

All the tissue samples were cut into small pieces (5*5*5mm^3^) and were stored at -80C in RNAlater solution until RNA was isolated.

#### 2. RNA isolation

Total RNA was isolated from Tissue using TRIzol reagent as per manufacturer protocol. Briefly, tissue was washed in sterile PBS to clean blood, bile etc. Visibly necrotic areas were dissected off. Approximately 5*5*5 mm3 tissue was homogenized in 1 ml of TRIzol reagent (2-Mercaptoethanol added).

From cell line total RNA was isolated using RNA isolation kit (Promega, Madison, USA).

#### 3. Nanostring nCounter analysis

Nanostring nCounter analysis was done with a customized panel for nuclear receptors according to the manufacturer’s protocol. Details of the probes are in Table S1.

#### 4. Realtime PCR

Realtime PCR was done (SYBR green) using specific primers (Table below). Agilent AriaMX Real time PCR system (Agilent Technologies, Santa clara, USA) was used for the study.

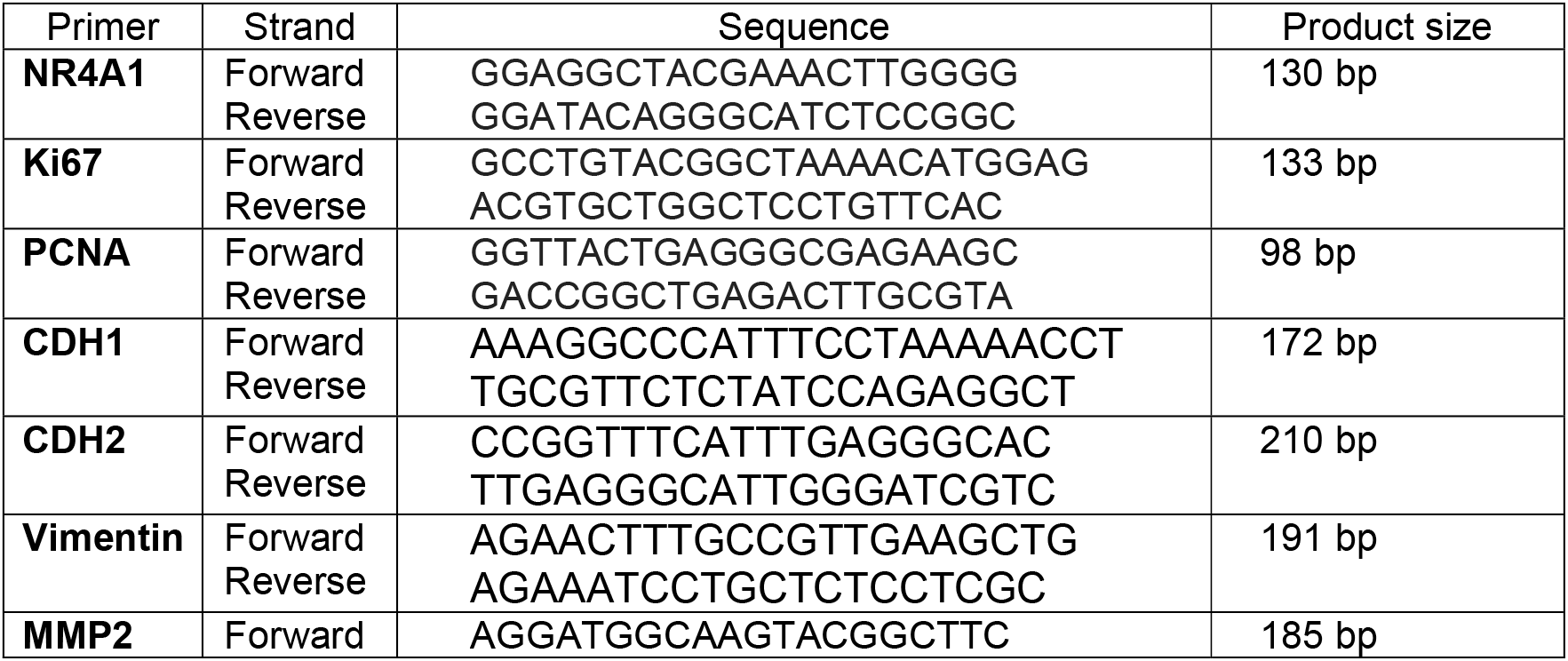
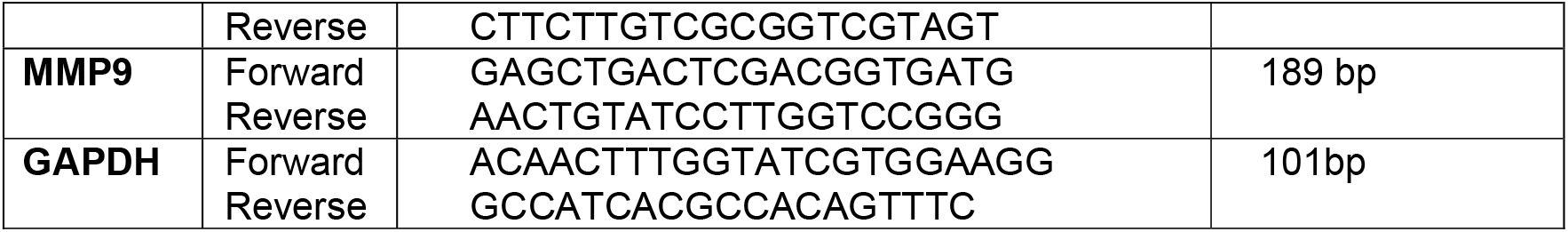

#### 5. Detection of protein by Western Blotting

Cells or tissue homogenized in ice-cold cell lysis buffer RIPA containing protease and phosphatase inhibitor cocktail. Protein concentrations were determined by the BCA assay (Pierce, Rockford, IL, USA). Equal amounts of protein (30 μg) were separated on SDS/10% PAGE, transferred onto Nitrocellulose membranes and immunoblotted overnight with anti-NR4A1 (Novusbio NB100-56745) or anti-GAPDH antibody (Affinity Biotech-AF0911). Membranes were washed in Tris-buffered saline (TBS)-0.1% Tween 20 solution prior to incubation with antiRabbit secondary horseradish peroxidase (HRP)-conjugated antibody (Cell Signal Technology-7074). HRP activity was revealed by incubation with the ECL substrate. Chemiluminescence reactions were visualized by Azure 200 (Azure Biosystems, Dublin, US). Images were quantitatively analyzed using ImageJ software (National Institute of Health, Bethesda, US). The membranes were stripped off all antibodies by means of incubating in stripping buffer without beta-mercaptoethanol followed by processing for housekeeping/loading control GAPDH.

#### 6. Cell culture characterization and treatment/ Knock-down

GBC cell lines NOZ and TGBC24TKB were a generous gift from IOB, Bengaluru, India. Cells were maintained in DMEM high glucose supplemented with 10% FBS and 1% antibiotic-antimycotic (Himedia, Thane, India). Cells were serum starved for 6 hours before treatment with CSNB 10uM (MedChemExpress, New Jersey, US) in DMEM containing 5%FBS. For knockdown experiments, 0.3 million cells were plated in a well of 6 well plate with NR4A1 siRNA (Thermo Fisher Scientific, Massachusetts, US) in lipofectamine-optiMEM (Invitrogen, Carlsbad, US) (final concentration of siRNA 20nM).

#### 7. Propidium iodide staining for Cell cycle

Cells were harvested by trypsinization following different treatments and washed twice with PBS. Fixation was done by adding cold 70% ethanol drop-wise to the cell pellet while vortexing. And was kept for 30 minutes at 4°C. They were washed 2 times in PBS, spun at 850 g in a centrifuge. The cells were treated with ribonuclease by adding 50 µl of a 100 µg/ml stock of RNase. 200 µl PI was added from a 50 µg/ml stock solution. flow cytometric analysis was done by LSR Fortessa (Becton, Dickinson and Company, New Jersy, US) using appropriate gating.

#### 8. Migration by wound healing assay

Cells were grown to 100% confluency in 35mm petri dishes. A uniform wound was created by a blunt tip to minimize damage to the adjacent cells. Appropriate treatment was administered after washing twice with sterile PBS. Serial images were taken at 10-15 min interval by Cytosmart Lux2 imaging system (Axion Biosystems, Atlanta, US) until the control wound healed. A timelapse video was created for each experiment.

#### 9. Transwell Matrigel Invasion assay

Eight-micron transwell inserts were coated with Matrigel (Corning, New York, USA) after 1:8 dilution with serum free media. 30 thousand cells were plated in a transwell and allowed to adhere overnight. Next day media of the lower chamber was replaced with serum free one thus creating a serum gradient. Specific treatments were added in the upper chamber. After overnight incubation the membranes were harvested fixed with chilled methanol and stained with crystal violet. The migrated cells on the bottom surface were counted using an inverted microscope.

#### 10. Statistical analysis

All the experiments were done at least 3 times with at least 2 technical replicates. Mann-Whitney U test or student t test was done to compare between 2 groups while one way ANOVA or Kruskal-Wallis test was done to compare among 3 or more groups with appropriate post hoc test. Graphpad Prism version 9 was used for statistical analysis and plotting the graphs.

