## Supplementary data for "High throughput screening of nuclear receptors identifies NR4A1, a novel tumor suppressor with potential as a therapeutic target in gallbladder cancer"

### Supplementary Table S1

Nanostring nCounter probe list

| | Customer Identifier | Accession | Position | Target Sequence | Flags | HUGO Gene |
|---|---|---|---|---|---|---|
| 1 | GUSB | NM_000181.3 | 1900-1999 | CCGATTTCATGACTGAACAGTCACCGACGAGAGTGCTGGGGAATAAAAAGGGATCTTCACTCGGCAGAGACAACCAAAAAGTGCAGCGTTCCTTTTGCG | HK | GUSB |
| 2 | HPRT1 | NM_000194.1 | 241-340 | TGTGATGAAGGAGATGGGAGGCCATCACATTGTAGCCCTCTGTGTGCTCAAGGGGGGCTATAAATTCTTTGCTGACCTGCTGGATTACATCAAAGCACTG | HK | HPRT1 |
| 3 | PGK1 | NM_000291.2 | 1031-1130 | GCAAGAAGTATGCTGAGGCTGTCACTCGGGCTAAGCAGATTGTGTGGAATGGTCCTGTGGGGGTATTTGAATGGAAGCTTTTGCCCGGGGAACCAAAGC | HK | PGK1 |
| 4 | TBP | NM_001172085.1 | 588-687 | ACAGTGAATCTTGGTTGTAAACTTGACCTAAAGACCATTGCACTTCGTGCCCGAAACGCCGAATATAATCCCAAGCGGTTTGCTGCGGTAATCATGAGGA | HK | TBP |
| 5 | TUBB | NM_178014.2 | 1956-2055 | TTCTAAGTATGTCCATTTCCCATCTCAGCTTCAAGGGAGGTGTCAGCAGTATTATCTCCACTTTCAATCTCCCTCCAAGCTCTACTCTGGAGGAGTCTGT | HK | TUBB |
| 6 | ACTB | NM_001101.2 | 1011-1110 | TGCAGAAGGAGATCACTGCCCTGGCACCCAGCACAATGAAGATCAAGATCATTGCTCCTCCTGAGCGCAAGTACTCCGTGTGGATCGGCGGCTCCATCCT | HK;X | ACTB |
| 7 | AR | NM_000044.2 | 876-975 | AGACTGTGAGCCTAGCAGGGCAGATCTTGTCCACCGTGTGTCTTCTTCTGCACGAGACTTTGAGGCTGTCAGAGCGCTTTTTGCGTGGTTGCTCCCGCAA | | AR |
| 8 | ESR1 | NM_000125.2 | 1596-1695 | AGGAACCAGGGAAAATGTGTAGAGGGCATGGTGGAGATCTTCGACATGCTGCTGGCTACATCATCTCGGTTCCGCATGATGAATCTGCAGGGAGAGGAGT | | ESR1 |
| 9 | ESR2 | NM_001214903.1 | 167-266 | AGCACGGCTCCATATACATACCTTCCTCCTATGTAGACAGCCACCATGAATATCCAGCCATGACATTCTATAGCCCTGCTGTGATGAATTACAGCATTCC | | ESR2 |
| 10 | ESRRA | NM_004451.3 | 856-955 | ACCCTCTGTGACCTCTTTGACCGAGAGATTGTGGTCACCATCAGCTGGGCCAAGAGCATCCCAGGCTTCTCATCGCTGTCGCTGTCTGACCAGATGTCAG | | ESRRA |
| 11 | ESRRB | NM_004452.3 | 953-1052 | CCCATACCTGAGCTTACAAATTTCTCCACCTGCTAAAAAGCCATTGACCAAGATTGTCTCATACCTACTGGTGGCTGAGCCGGACAAGCTCTATGCCATG | | ESRRB |
| 12 | ESRRG | NM_001134285.1 | 2848-2947 | CTGACACAAAGGATGAGATTACACCTACAGTGACTCTGAGTAGTCACATATATAAGCACTGCACATGAGATATAGATCCGTAGAATTGTCAGGAGTGCAC | | ESRRG |
| 13 | HNF4A | NM_178850.1 | 1117-1216 | CCTTGCAGAGCATCACCTGGCAGATGATCGAGCAGATCCAGTTCATCAAGCTCTTCGGCATGGCCAAGATTGACAACCTGTTGCAGGAGATGCTGCTGGG | | HNF4A |
| 14 | HNF4G | NM_004133.3 | 2196-2295 | CTATAGGTGGAAAGTCTGTGTCAGCCAAAGCCATGTCACTCTGAGAGAACTGATTCTGAGGACAAGTTAGCCATCTACCACCCATGTCCCTTATTTGGTC | | HNF4G |
| 15 | NR0B1 | NM_000475.4 | 1346-1445 | TTGCTGAACTGTTCTTCAGGCCCATCATCGGCACAGTCAGCATGGATGATATGATGCTGGAAATGCTCTGTACAAAGATATAAAGTCATGTGGGCCACAC | | NR0B1 |
| 16 | NR0B2 | NM_021969.1 | 736-835 | TCTTTCGCCCTATCATTGGAGATGTTGACATCGCTGGCCTTCTTGGGGACATGCTTTTGCTCAGGTGACCTGTTCCAGCCCAGGCAGAGATCAGGTGGGC | | NR0B2 |
| 17 | NR1D1 | NM_021724.3 | 1081-1180 | GCAAGGGCTTTTTCCGTCGGAGCATCCAGCAGAACATCCAGTACAAAAGGTGTCTGAAGAATGAGAATTGCTCCATCGTCCGCATCAATCGCAACCGCTG | | NR1D1 |
| 18 | NR1D2 | NM_001145425.1 | 2001-2100 | AGCTGTTCGGATTGAGAACTCTTCAGCCATGATTAGACGTTGACTGCATCTCCCTGATAGACCAATCAGCTGTGTCGCACTTAAACTGGAGAAGTTACAC | | NR1D2 |
| 19 | NR1H2 | NM_007121.4 | 31-130 | TGACGCAAGGGCGGGGTTGCCGGAAGAAGTGGCGAAGTTACTTTTGAGGGTATTTGAGTAGCGGCGGTGTGTCAGGGGCTAAAGAGGAGGACGAAGAAAA | | NR1H2 |
| 20 | NR1H3 | NM_005693.2 | 1576-1675 | CCCATATTTTCTGTTTTCTTGGCCGGATGGCTGAGGCCTGGTGGCTGCCTCCTAGAAGTGGAACAGACTGAGAAGGGCAAACATTCCTGGGAGCTGGGCA | | NR1H3 |
| 21 | NR1H4 | NM_001206977.1 | 985-1084 | GGGCGCGTCAGCAGGGAGGATCAAAGGGGATGAGCTGTGTGTTGTTTGTGGAGACAGAGCCTCTGGATACCACTATAATGCACTGACCTGTGAGGGGTGT | | NR1H4 |
| 22 | NR1H5P | ENST00000452683.1 | 846-945 | TCAGGAAAGCATAGCTGCAGGAACATACAAATCCTGAACTGAGCTTTTTGCAACTCTCAGAGACAGCAGTCCTACACATACGTGGGCTAATGAATTTTAC | | NR1H5P |
| 23 | NR1I2 | NM_022002.2 | 1266-1365 | GCAATTCGCCATTACTCTGAAGTCCTACATTGAATGCAATCGGCCCCAGCCTGCTCATAGGTTCTTGTTCCTGAAGATCATGGCTATGCTCACCGAGCTC | | NR1I2 |
| 24 | NR1I3 | NM_005122.3 | 661-760 | CTGTGCTGCCTCTGGTCACACACTTCGCAGACATCAACACTTTCATGGTACTGCAAGTCATCAAGTTTACTAAGGACCTGCCCGTCTTCCGTTCCCTGCC | | NR1I3 |
| 25 | NR2C1 | NM_003297.2 | 336-435 | AGAAAATCCAGATTGTGACAGCACTTGATCATAATACCCAAGGCAAGCAGTTCATTCTGACAAATCACGACGGCTCTACTCCAAGCAAAGTCATTCTGGC | | NR2C1 |
| 26 | NR2C2 | NM_003298.3 | 1795-1894 | TTTGACCAGCACAAGCCAGATTGAAAAATTCCAAGAAAAGGCACAGATGGAGTTGCAGGACTATGTTCAGAAAACCTACTCAGAAGACACCTACCGATTG | | NR2C2 |
| 27 | NR2E1 | NM_003269.3 | 1531-1630 | ACAAGATCATATCTGAAATACAGGCTTTACAAGAGGTGGTGGCTCGATTTAGACAACTCCGGTTAGATGCTACTGAATTTGCCTGTCTAAAATGCATCGT | | NR2E1 |
| 28 | NR2E3 | NM_014249.2 | 859-958 | CTCGGCTCGCCTACTCTTCATGGCCGTCAAGTGGGCCAAGAACCTGCCTGTGTTCTCCAGCCTGCCCTTCCGGGATCAGGTGATCCTGCTGGAAGAGGCG | | NR2E3 |
| 29 | NR2F1 | NM_005654.4 | 3111-3210 | AGCAGAAATACAATCCGAGCTACAAAGCATGGGAAAAAGAGACTCTTTTAGGATCAGATCTGTGAGCACGTTGGCGAGGAAAAACAAAAAAAAAAAAAA | | NR2F1 |
| 30 | NR2F2 | NM_021005.2 | 1531-1630 | CCATAGTCCTGTTCACCTCAGATGCCTGTGGTCTCTCTGATGTAGCCCATGTGGAAAGCTTGCAGGAAAAGTCTCAGTGTGCTTTGGAAGAATACGTTAG | | NR2F2 |
| 31 | NR2F6 | NM_005234.3 | 1695-1794 | CAGTGGGCAGGGGTAGCTAGCGTGTGCCAGGCAGATCCTCTGGACACGTAACCTATGTCAGACACTACATGATGACTCAAGGCCAATAATAAAGACATTT | | NR2F6 |
| 32 | NR3C1 | NM_001018077.1 | 2823-2922 | GGAGATCATATAGACAATCAAGTGCAAACCTGCTGTGTTTTGCTCCTGATCTGATTATTAATGAGCAGAGAATGACTCTACCCTGCATGTACGACCAATG | | NR3C1 |
| 33 | NR3C2 | NM_000901.3 | 431-530 | CCTACAGAGAGGACCGATGAGAATAACTACATGGAGATTGTCAACGTAAGCTGTGTTTCCGGTGCTATTCCAAACAACAGTACTCAAGGAAGCAGCAAAG | | NR3C2 |
| 34 | NR4A1 | NM_173157.1 | 1576-1675 | ACCGGCTGCAGTGTGCCCGTGGCTTCGGGGACTGGATTGACAGTATCCTGGCCTTCTCAAGGTCCCTGCACAGCTTGCTTGTCGATGTCCCTGCCTTCGC | | NR4A1 |
| 35 | NR4A2 | NM_006186.3 | 1381-1480 | TTCAGAAGTGCCTGGCTGTTGGGATGGTCAAAGAAGTGGTTCGCACAGACAGTTTAAAAGGCCGGAGAGGTCGTTTGCCCTCGAAACCGAAGAGCCCACA | | NR4A2 |
| 36 | NR4A3 | NM_006981.2 | 1841-1940 | GAGGTCGTCTGCCTTCCAAACCAAAGAGCCCATTACAACAGGAACCTTCTCAGCCCTCTCCACCTTCTCCTCCAATCTGCATGATGAATGCCCTTGTCCG | | NR4A3 |
| 37 | NR5A1 | NM_004959.4 | 2186-2285 | CTCTGCAAGAGGGGGCATTGATACATCATCGGGAAAAAACTTTGCTCCAGGCATCACTGATTCCCTCTCCCACCCAAGGAGAACGTTTGGTACAATCGAC | | NR5A1 |
| 38 | NR5A2 | NM_003822.3 | 3821-3920 | ATGAAGGGTACACATTAGGTAAGCTGGGCGTTGACTCATGCGCAGTCTCAGTCACCCGTGTTATCTTCGTGGCTCAAAGGACAATGCAAAATCGCCGATC | | NR5A2 |
| 39 | NR6A1 | NM_033335.2 | 971-1070 | GCCGACGAGCTGCTCTTTAGGCAGATTGCCTGGATCAAGAAACTGCCTTTCTTCTGCGAGCTCTCAATCAAGGATTACACGTGCCTCTTGAGCTCTACGT | | NR6A1 |
| 40 | PGR | NM_000926.4 | 2393-2492 | AGCCAGCCAGAGCCCACAATACAGCTTCGAGTCATTACCTCAGAAGATTTGTTTAATCTGTGGGGATGAAGCATCAGGCTGTCATTATGGTGTCCTTACC | | PGR |
| 41 | PPARA | NM_001001928.2 | 267-366 | CAAGAAATGGGAAACATCCAAGAGATTTCGCAATCCATCGGCGAGGATAGTTCTGGAAGCTTTGGCTTTACGGAATACCAGTATTTAGGAAGCTGTCCTG | | PPARA |
| 42 | PPARD | NM_006238.4 | 896-995 | AGGCCCGCAGCATCCTCACCGGCAAAGCCAGCCACACGGCGCCCTTTGTGATCCACGACATCGAGACATTGTGGCAGGCAGAGAAGGGGCTGGTGTGGAA | | PPARD |
| 43 | PPARG | NM_005037.5 | 346-445 | CAGATCCAGTGGTTGCAGATTACAAGTATGACCTGAAACTTCAAGAGTACCAAAGTGCAATCAAAGTGGAGCCTGCATCTCCACCTTATTATTCTGAGAA | | PPARG |
| 44 | RARA | NM_001024809.3 | 1306-1405 | ACAGCTCAGAACAACGTGTCTCTCTGGACATTGACCTCTGGGACAAGTTCAGTGAACTCTCCACCAAGTGCATCATTAAGACTGTGGAGTTCGCCAAGCA | | RARA |
| 45 | RARB | NM_000965.3 | 2756-2855 | TAACAACTCCCAAAGAAACAGGCATAGAATCTGCCTCCTTTGACCTTGTTCAATCACTATGAAGCAGAGTGAAAGCTGTGGTAGAGTGGTTAACAGATAC | | RARB |
| 46 | RARG | NM_000966.3 | 1541-1640 | GCCAGCCCTACATGTTCCCAAGGATGCTAATGAAAATCACCGACCTCCGGGGCATCAGCACTAAGGGAGCTGAAAGGGCCATTACTCTGAAGATGGAGAT | | RARG |
| 47 | RORA | NM_134261.2 | 1716-1815 | AAAATTAACCGAGACACTTTATATGGCCCTGCACAGACCTGGAGCGCCACACACTGCACATCTTTTGGTGATCGGGGTCAGGCAAAGGAGGGGAAACAAT | | RORA |
| 48 | RORB | NM_006914.3 | 1841-1940 | CCTGGATGATGAGACCTTGGCAAAGTTAATAGCCAAGATACCAACCATCACGGCAGTTTGCAACTTGCACGGGGAGAAGCTGCAGGTATTTAAGCAATCT | | RORB |
| 49 | RORC | NM_001001523.1 | 1351-1450 | CTCATCAATGCCCATCGGCCAGGGCTCCAAGAGAAAAGGAAAGTAGAACAGCTGCAGTACAATCTGGAGCTGGCCTTTCATCATCATCTCTGCAAGACTC | | RORC |
| 50 | RXRA | NM_002957.4 | 5051-5150 | TACAAATGTAATTTTATCCCTCATGTATACTTGGATATGGCGGGGGGAGGGCTGGGACTGTTTCGTTTCTGCTTCTAGAGATTGAGGTGAAAGCTTCGTC | | RXRA |
| 51 | RXRB | NM_021976.3 | 1251-1350 | CCACACTTTTCCTCCTTGCCTCTGGATGATCAGGTCATATTGCTGCGGGCAGGCTGGAATGAACTCCTCATTGCCTCCTTCTCACACCGATCCATTGATG | | RXRB |
| 52 | RXRG | NM_006917.3 | 1106-1205 | TATGTCATGCTGCTGACAAGCAGCTTTTCACCCTCGTTGAATGGGCCAAGCGTATTCCCCACTTCTCTGACCTCACCTTGGAGGACCAGGTCATTTTGCT | | RXRG |
| 53 | THRA | NM_199334.2 | 651-750 | GAAAGCGAAAAAGAAAGAACGGCCAATGTTCCCTGAAAACCAGCATGTCAGGGTATATCCCTAGTTACCTGGACAAAGACGAGCAGTGTGTCGTGTGTGG | | THRA |
| 54 | THRB | NM_000461.4 | 386-485 | AAGCCTGCCTACATAGGAAGAGCCATTCAGAGAGGCGCAGCACGTTGAAAAATGAACAGTCGTCGCCACATCTCATCCAGACCACTTGGACTAGCTCAAT | | THRB |
| 55 | VDR | NM_000376.2 | 984-1083 | CCAATGAGTCCTTCACCATGGACGACATGTCCTGGACCTGTGGCAACCAAGACTACAAGTACCGCGTCAGTGACGTGACCAAAGCCGGACACAGCCTGGA | | VDR |

### Supplementary Table S2

Report of STR profiling of the GBC cell lines used done by the authors compared to that of the cell repository.

| Cell line | NOZ | |
|---|---|---|
| Loci | Database(JCRB) | STR profile done by NCCS |
| Amelogenin | X | X |
| CSF1PO | 11,13 | 11,13 |
| D5S818 | 11,13 | 11,13 |
| D7S820 | 10,11 | 10,11 |
| D13S317 | 8,12 | 8,12 |
| D16S539 | 9,11 | 9,11 |
| TH01 | 7,9 | 7,9 |
| TPOX | 8,11 | 8,11 |
| vWA | 19 | 19 |

| Cell line | TGBC24TKB | |
|---|---|---|
| Loci | Database(JCRB) | STR profile done by NCCS |
| Amelogenin | X | X |
| CSF1PO | 12,13 | 12,13 |
| D5S818 | 11 | 11 |
| D7S820 | 10,11 | 10,11 |
| D13S317 | 12 | 12 |
| D16S539 | 9,10 | 9,10 |
| TH01 | 6 | 6 |
| TPOX | 8 | 8 |
| vWA | 16 | 16 |

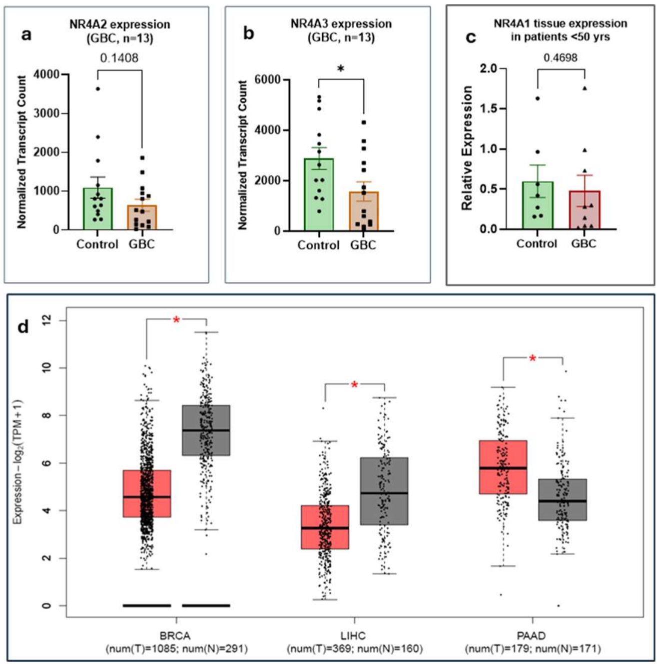


**Supplementary Figure S1:** a,b) mRNA expression of 2 other members of NR4A family NR4A2 and NR4A3 respectively, by Nanostring nCounter. c) mRNA expression of NR4A1 in patients with age <50years had no significant difference with that of the age matched controls. d)TCGA database shows both downregulation and upregulation of NR4A1 are observed in different cancers.

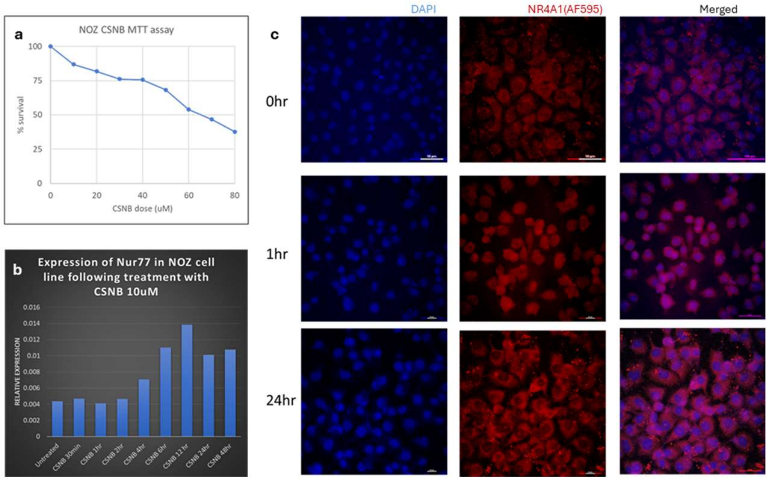


**Supplementary Figure S2:** Validation of CSNB as an agonist of NR4A1. a) MTT assay shows the $LD_{50}$ around 60μM. b) mRNA expression of NR4A1/Nur77 following CSNB treatment is time dependent. c) Nuclear translocation of the receptor following agonist treatment.

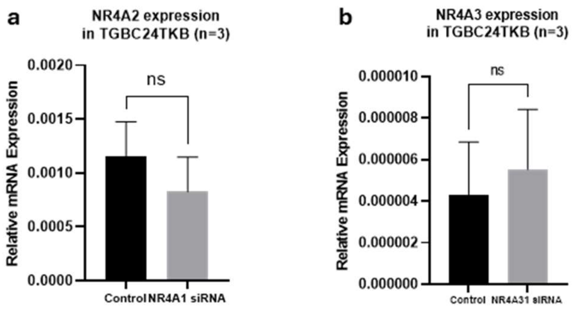


**Supplementary Figure S3:** Expression of the other members of NR4A family is not altered significantly following siRNA mediated knockdown of NR4A1.
